# Cross-Tabulating Epidemiological Covariates with AUDIT-C Data in Large-Scale Biobanks

**DOI:** 10.64898/2026.04.01.26349975

**Authors:** August Blackburn

## Abstract

**Introduction:** The Alcohol Use Disorders Identification Test-Consumption (AUDIT-C) is a widely utilized screening tool in large-scale electronic health record (EHR) biobanks. However, its categorical, range-based survey responses present a significant challenge for epidemiological research, especially where continuous quantitative variables may be preferred. Standard workarounds, such as assigning categorical midpoints or utilizing aggregate ordinal scores for regression mapping often introduce false mathematical precision or obscure critical behavioral nuances between drinking frequency and quantity. This report presents a novel framework for presenting and bounding categorical alcohol survey data.

**Materials and Methods:** I developed two complementary descriptive techniques: (1) a two-dimensional cross-tabulation matrix that preserves the interaction between drinking frequency and typical quantity, and (2) a systematic bounding algorithm that applies time-interval correction factors to calculate strict lower and upper estimates of average daily alcohol consumption. To demonstrate the real-world utility of this framework, I applied these methods to three analytical descriptive scenarios within a European ancestry (EUR) cohort of the *All of Us* Research Program: Generalized Anxiety Disorder (GAD) prevalence (n=104,893), minor allele frequency (MAF) for the rs1229984 genetic variant (n=104,890), and self-reported active duty military service history (n=104,893).

**Results:** Application of the cross-tabulation matrix revealed patterns across all three descriptive scenarios. For example, participants reporting the highest frequency (“4 or more times a week”) combined with the highest quantity (“10 or More” drinks) demonstrated a GAD prevalence of 13.5%, compared to 5.8% among those reporting the same frequency but a low quantity (“1 or 2” drinks). A general trend of increased anxiety in higher quantity drinkers contrasts with a general trend of decreased anxiety in higher frequency drinkers. Bounding estimates for average daily consumption ranged from 0.299 to 0.730 drinks for individuals with GAD, and 0.303 to 0.787 for those without. Those who reported having been active duty in the US Armed Forces demonstrated a general trend toward more frequent drinking and higher average daily consumption estimates (0.339 to 0.875) than those who had not (0.297 to 0.770). The minor allele of the genetic variant rs1229984 exhibited a clear effect reducing both frequency and quantity, resulting in lower average daily consumption estimates.

**Conclusions:** This bounding and mapping framework provides researchers with an additional method to traditional midpoint and aggregate scoring methods. By explicitly defining the uncertainty inherent in categorical survey instruments and visualizing cohort distributions across intersecting behavioral axes, this methodology improves the resolution, reproducibility, and interpretability of lifestyle exposure data.

## Introduction

The proliferation of large-scale, EHR-linked biobanks, such as the National Institutes of Health *All of Us* Research Program^1^, has provided researchers with unprecedented opportunities to conduct highly powered epidemiological and genetic studies. However, while clinical outcomes can often be objectively defined using standardized billing codes, critical lifestyle covariates such as alcohol consumption are predominantly captured through self-reported surveys. Instruments like the Alcohol Use Disorders Identification Test-Consumption^2^ are widely utilized in these cohorts due to their validated clinical utility and low respondent burden. This is particularly relevant in military and veteran populations, where the AUDIT-C is routinely administered across the Veterans Health Administration (VHA) and Department of Defense (DoD).^3-5^

Despite their widespread use, surveys like the AUDIT-C present a distinct methodological challenge for quantitative research: they capture continuous behaviors using categorical, range-based bins. For example, participants report their drinking frequency (e.g., “2 to 4 times a month”) and typical drinking quantity (e.g., “3 or 4 drinks”) by selecting from pre-defined qualitative options. While this categorical approach works well for its intended use case (primary screening)^2^, it creates a significant hurdle for researchers because of the ordinal, semi-quantitative nature of the data.

Historically, researchers have circumvented the limitations of categorical survey data through two primary analytical workarounds. The most ubiquitous approach involves assigning arbitrary midpoints to the categorical bins (e.g., converting “3 or 4 drinks” to exactly 3.5) to force the calculation of a single, continuous daily ethanol exposure metric.^6,7^ However, this assigns exact values to range-based choices, generating an implied precision that may exceed the survey’s original design. Indeed, studies comparing the categorical AUDIT-C with continuous drinking measures have demonstrated that converting these categorical choices into exact continuous values obscures substantial variability in actual alcohol consumption.^8^ Alternatively, some investigators utilize regression mapping to map the aggregate ordinal AUDIT-C score (0-12) directly to a mean daily drinking estimate^9^. While highly effective for general screening, relying on the aggregate score may not capture the full variation of underlying behaviors; an identical score (and therefore an identical daily volume estimate) can be generated by a frequent light drinker and an infrequent binge drinker.

To complement these established methods, this methodological report presents a structured approach for organizing, presenting, and contextualizing categorical AUDIT-C survey responses. Specifically, I detail two descriptive techniques: (1) a systematic bounding approach that applies time-interval correction factors to cross-tabulated survey responses, providing theoretical ‘low’ and ‘high’ estimates for average daily alcohol consumption to help appropriately scale and contextualize the categorical bins, and (2) a standardized cross-tabulation matrix designed to clearly present cohort distributions and clinical outcome prevalences across these intersecting behavioral strata. By detailing this presentation strategy, this paper aims to provide researchers with additional tools to report alcohol-related survey data and summarize complex behavioral phenotypes within large-scale observational cohorts.

## Methods

### Demonstration Cohorts

The methods detailed in this report were developed during, and applied to, specific analytical subsets of the *All of Us* Research Program curated for primary epidemiological investigations.^10^ Rather than generating a new cohort, utilizing these exact datasets allows for a transparent methodological companion detailing how complex categorical exposure variables can be mapped and contextualized. To demonstrate the versatility of this framework, it was applied to three distinct domains:

#### Clinical Phenotyping

Evaluating Generalized Anxiety Disorder (GAD) prevalence within a European (EUR) ancestry cohort (n=104,893).

#### Genetic Epidemiology

Mapping the minor allele frequency (MAF) of the rs1229984 genetic variant across the same EUR cohort. This variant is a well-characterized missense mutation in the *ADH1B* gene. For this analysis, the sample size was restricted to 104,890 individuals; 3 participants were excluded because they carried a separate, very rare third allele at this locus.

#### Demographic Assessment

Evaluating alcohol consumption patterns based on self-reported active duty military service history (n=104,893). Specifically, participants were classified based on their response to the survey question: “Have you ever served on active duty in the US Armed Forces?”.

### Bounding Estimation for Average Daily Consumption

To provide a tangible, real-world context for the categorical AUDIT-C survey responses, we developed a bounding approach that estimates a theoretical range of average daily alcohol consumption. Rather than assigning a single arbitrary midpoint to each category, this method calculates lower and upper limits based on the cross-tabulation of reported drinking frequency and typical drinking quantity.

The standard AUDIT-C survey captures drinking frequency (*f*) and typical quantity (*q*) using categorical bins. For bounded categories (e.g., frequency of “2 to 4 times a month” or quantity of “3 or 4 drinks”), the discrete numerical minimums (*f*_*low*_, *q*_*low*_) and maximums (*f*_*high*_, *q*_*high*_) are extracted directly from the survey text. For open-ended categories (e.g., frequency of “Monthly or less” or “4 or more times a week”), reasonable absolute limits must be defined by the investigator based on the study population (e.g., anchoring “Monthly or less” to a frequency of exactly 1 time per month). In this manuscript I use the following choices for open-ended categories: a frequency of exactly 1 time per month for both low and high estimates for “Monthly or less”, 4 and 7 times per week for low and high estimates for “4 or More Per Week”, and 10 drinks for low and high drinks for “10 or more” daily drinks.

Once the numerical limits for frequency and quantity are defined for a given cross-tabulated category, the estimated average daily consumption (*E*) is calculated by multiplying the frequency by the quantity and dividing by a time-interval correction factor (*t*). The correction factor normalizes the reported timeframe into a daily rate (e.g., *t* = 7 for weekly frequencies, and *t* = 30.4375 for monthly frequencies to account for the average length of a month over a 365.25-day year).

The lower and upper bounds of average daily consumption for any specific survey response combination are computed using the following formulas:

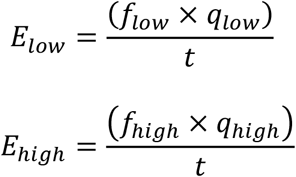

For example, for participants reporting a frequency of “monthly or less” *f*_*low*_ = 1, *f*_*high*_ = 1, *t* = 30.4375 and a typical quantity of “1 or 2 drinks” *q*_*low*_ = 1, *q*_*high*_ = 2, the estimated daily consumption range is calculated as 0.0329 (*E*_*low*_) to 0.0657 (*E*_*high*_) drinks per day. This limit-based calculation is systematically applied across all intersecting categories to generate a standardized matrix of consumption bounds.

### Cross-Tabulation Mapping Matrix

To accurately map cohort distributions and clinical outcome prevalences across these complex behavioral phenotypes, I utilized a two-dimensional cross-tabulation matrix. This presentation framework moves beyond unidimensional summaries to visualize the distinct interactions between drinking frequency and quantity.

### Matrix Construction

The matrix is structured with past-year drinking frequency serving as the rows and typical drinking quantity serving as the columns.

### Handling Incomplete Data

An optional component of this matrix is the inclusion of an “Unspecified” or “Prefer Not to Answer” column. Because survey logic often permits participants to skip questions, restricting the analysis only to complete cases could introduce selection bias. The “Unspecified” column retains these individuals in the denominator for frequency-level summary statistics. Here, I have discarded data for individuals who did not complete the full survey.

### Proportion Calculations

Within each intersecting cell of the matrix, the baseline prevalence (proportion) of a given clinical outcome is calculated by dividing the number of individuals with the target Electronic Health Record (EHR) diagnosis by the total number of individuals occupying that specific intersecting cell.

## Results

To demonstrate the broad utility of this approach across varying epidemiological domains, the cross-tabulation matrix and bounding algorithm were applied to three distinct analytical use cases.

### Application 1: Clinical Phenotyping (Generalized Anxiety Disorder)

When mapping GAD prevalence within the EUR cohort (n=104,893), the multidimensional matrix (**Table 1**) immediately highlights nuanced patterns that might be obscured by a single aggregate AUDIT-C score. The matrix reveals that individuals reporting the highest drinking frequency (“4 or more times a week”) combined with the highest quantity (“10 or More” drinks) had a GAD prevalence of 13.5%. Conversely, those reporting the same high frequency but a low quantity (“1 or 2” drinks) demonstrated a lower GAD prevalence of 5.8%. In the cross-tabulation matrix two contrasting trends are apparent: a general trend of higher anxiety rates in higher volume drinkers and a general trend of lower anxiety rates in higher frequency drinkers.

**Table 1.**

| Percent w/<br>Generalized Anxiety<br>Disorder Diagnoses<br>in EUR (n=104,893) |  | “On a typical day when you drink, how many drinks do<br>you have?” |  |  |  |  |  | All in<br>row |
| --- | --- | --- | --- | --- | --- | --- | --- | --- |
|  |  |  | 1 or 2 | 3 or 4 | 5 or 6 | 7 to 9 | 10 or<br>More |  |
| “How<br>often did<br>you have a<br>drink<br>containing<br>alcohol in<br>the past<br>year?” | Never | 0.095 |  |  |  |  |  | 0.095 |
|  | Monthly<br>or Less |  | 0.093 | 0.118 | 0.101 | 0.122 | 0.088 | 0.095 |
|  | 2 to 4<br>time a<br>month |  | 0.074 | 0.088 | 0.083 | 0.133 | 0.161 | 0.077 |
|  | 2 to 3<br>times a<br>week |  | 0.069 | 0.090 | 0.105 | 0.119 | 0.069 | 0.075 |
|  | 4 or<br>more<br>times a<br>week |  | 0.058 | 0.065 | 0.108 | 0.113 | 0.135 | 0.065 |
| All in column |  | 0.095 | 0.077 | 0.087 | 0.099 | 0.120 | 0.123 | 0.083 |

The bounding method estimates that individuals with a GAD diagnosis consumed between 0.299 (low estimate) and 0.730 (high estimate) drinks per day. Individuals without a GAD diagnosis consumed between 0.303 and 0.787 drinks per day. When calculating the ratio of average daily consumption for individuals with GAD versus those without, the ratio is 0.98 for the low estimate and 0.93 for the high estimate.

### Application 2: Genetic Epidemiology (rs1229984)

Applying this framework to genetic data allows for presentation of the suppressive effects of the rs1229934 minor allele on alcohol consumption. The cross-tabulation matrix for the rs1229984 genetic variant in the EUR cohort (n=104,890; **Table 2**) shows a distinctly lower minor allele frequency (MAF) at higher drinking frequencies and quantities, with an MAF of 0.010 in the highest frequency and quantity bin (“4 or more times a week” and “10 or More” drinks).

**Table 2.**
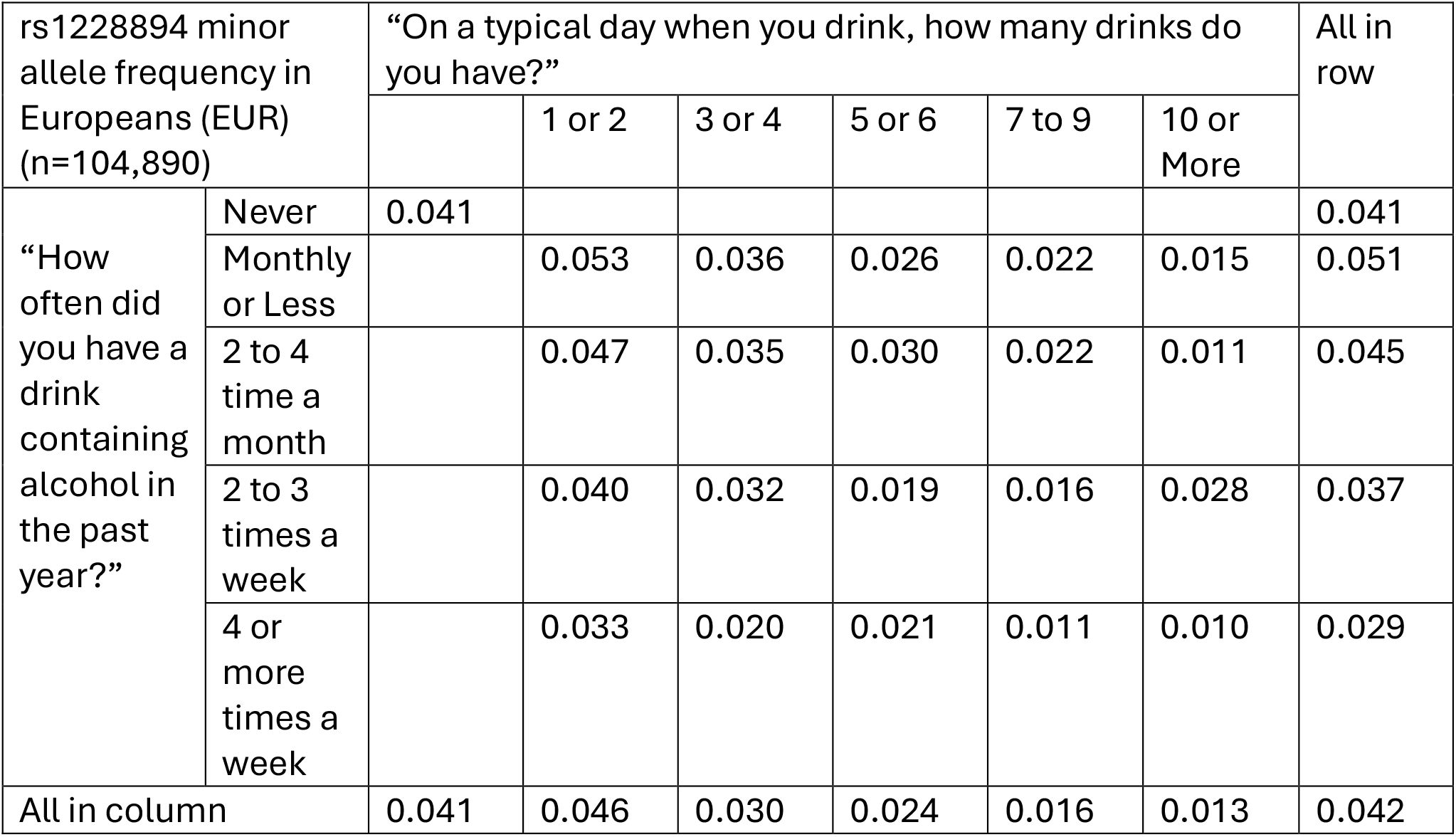

| rs1228894 minor allele frequency in Europeans (EUR) (n=104,890) |  | “On a typical day when you drink, how many drinks do you have?” |  |  |  |  |  | All in row |
| --- | --- | --- | --- | --- | --- | --- | --- | --- |
|  |  |  | 1 or 2 | 3 or 4 | 5 or 6 | 7 to 9 | 10 or More |  |
| “How often did you have a drink containing alcohol in the past year?” | Never | 0.041 |  |  |  |  |  | 0.041 |
|  | Monthly or Less |  | 0.053 | 0.036 | 0.026 | 0.022 | 0.015 | 0.051 |
|  | 2 to 4 time a month |  | 0.047 | 0.035 | 0.030 | 0.022 | 0.011 | 0.045 |
|  | 2 to 3 times a week |  | 0.040 | 0.032 | 0.019 | 0.016 | 0.028 | 0.037 |
|  | 4 or more times a week |  | 0.033 | 0.020 | 0.021 | 0.011 | 0.010 | 0.029 |
| All in column |  | 0.041 | 0.046 | 0.030 | 0.024 | 0.016 | 0.013 | 0.042 |

The bounding algorithm yields dose-dependent estimates of daily alcohol exposure based on genotype. Individuals harboring 0 copies of the minor allele had estimated consumption ranges of 0.311 to 0.803 drinks per day. This dropped to 0.201 to 0.552 for those with 1 copy, and 0.191 to 0.550 for those with 2 copies. This approach allows researchers to approximate the effectiveness of a genetic instrument. Individuals with 1 copy of the minor allele consumed 64.6-68.7% as much alcohol as those with 0 copies, based on the low and high boundary estimates. Individuals with 2 copies of the minor allele consumed 61.5-68.5% as much alcohol as those with 0 copies.

### Application 3: Demographic Assessment (Active Duty Military Service)

Finally, the matrix is highly effective for isolating behavioral differences across demographic variables. Among participants answering the question regarding active duty military service (n=104,915; **Table 3**), the matrix highlights an elevated concentration of those with active duty military service within the highest frequency drinking strata (“4 or more time per week”).

**Table 3.**

| Percent “Yes” on Active Duty Question (n=104,893) |  | “On a typical day when you drink, how many drinks do you have?” |  |  |  |  |  | All in row |
| --- | --- | --- | --- | --- | --- | --- | --- | --- |
|  |  |  | 1 or 2 | 3 or 4 | 5 or 6 | 7 to 9 | 10 or More |  |
| “How often did you have a drink containing alcohol in the past year?” | Never | 0.159 |  |  |  |  |  | 0.159 |
|  | Monthly or Less |  | 0.105 | 0.092 | 0.090 | 0.096 | 0.103 | 0.103 |
|  | 2 to 4 time a month |  | 0.104 | 0.090 | 0.088 | 0.094 | 0.161 | 0.101 |
|  | 2 to 3 times a week |  | 0.106 | 0.104 | 0.103 | 0.135 | 0.069 | 0.106 |
|  | 4 or more times a week |  | 0.143 | 0.160 | 0.181 | 0.172 | 0.130 | 0.149 |
| All in column |  | 0.159 | 0.111 | 0.114 | 0.121 | 0.133 | 0.123 | 0.121 |

The bounding algorithm confirms this shift in absolute volume, calculating that participants with active duty experience had approximately 1.14 times the alcohol consumption based on an estimated daily consumption range of 0.339 to 0.875 drinks per day, compared to a range of 0.297 to 0.770 drinks per day for those without active duty experience.

## Discussion

### Novelty and Methodological Advantages

While the fundamental calculation of multiplying drinking frequency by quantity is a cornerstone of alcohol epidemiology^11^, the framework presented in this report offers an additional approach to complement current data presentation practices. The primary utility lies in formalizing a two-dimensional descriptive mapping tool paired with a strict bounding algorithm, which together present an additional tool for analyzing alcohol consumption data from the AUDIT-C and similar ordinal semi-quantitative surveys.

Unlike regression mapping approaches that rely on aggregate scores and may group distinct behaviors together, this cross-tabulation matrix explicitly preserves the critical interaction between frequency and quantity. Furthermore, by calculating strict absolute bounds (the low and high estimates) rather than assigning arbitrary midpoints, this method transparently acknowledges and quantifies the uncertainty inherent in categorical survey instruments rather than implying mathematical precision that the survey was not designed to capture. This matrix provides a universally readable, purely descriptive tool that any clinician or researcher can interpret without needing to deconstruct an underlying statistical model. This transparency is particularly valuable for military medicine applications, allowing clinical administrators and researchers to visualize the precise behavioral distributions of active duty and veteran populations.

### Limitations

There are several limitations to this proposed framework. First, because the inputs rely on the AUDIT-C, the calculated bounds remain subject to standard self-report limitations and biases. Second, the bounding method requires investigators to make necessary assumptions when defining the absolute limits for open-ended categories (e.g., capping “10 or more drinks” at exactly 10). While this provides a mathematical foundation for analysis, variations in how these open-ended limits are defined across different studies could hinder direct cross-study comparability.

## Conclusion

These methods offer significant advantages for epidemiological and genetic research. By presenting consumption bounds rather than artificial point estimates, researchers can identify general trends and approximate daily alcohol consumption without overrepresenting the precision inherent in the data. Furthermore, the cross-tabulation matrix ensures that critical subpopulations and trends in frequency and quantity can be evaluated in the analysis. Ultimately, adopting this presentation strategy can improve the resolution, reproducibility, and interpretability of lifestyle exposure data in modern, large-scale biobanks.

## Data Availability

The data underlying this article cannot be shared publicly due to the privacy and security protocols of the National Institutes of Health. The data are available to registered researchers who are granted access to the secure All of Us Researcher Workbench cloud environment following institutional affiliation and data use agreements.

https://researchallofus.org/

## Declarations

Not applicable.

## Acknowledgements

The author gratefully acknowledge the participants of the National Institutes of Health *All of Us* Research Program for their indispensable contributions to this dataset, without whom this research would not have been possible. Additionally, during the preparation of this work, the author used Google Gemini to aid in manuscript drafting and language editing. The author independently verified all AI-assisted text, maintained full control over the scientific arguments, and bears ultimate responsibility for the manuscript.

## Prior presentation

Not applicable.

## Clinical Trial Registration

Not applicable

## Institutional Review Board (Human Subjects)

This study utilized de-identified, secondary data from the *All of Us* Research Program and was reviewed by the Air Force Research Laboratory Institutional Review Board, which determined the activity in the protocol is research but does not involve “human subjects” under the Common Rule (32 CFR 219) (Protocol: FWR2024132N v1.01).

## Institutional Animal Care and Use Committee (IACUC)

Not applicable

## Individual author contribution statements

AB conceived of and designed the research, performed all data analysis, and drafted the manuscript.

## Disclaimer

“The views expressed are those of the authors and do not reflect the official views or policy of the Department of Defense or its Components”

## Institutional clearance

Distro A: Cleared for Public Release, AFRL/PA, AFRL-2026-1119, 10 Mar 26

## References

1. All of Us Research Program Investigators. The “All of Us” Research Program. N Engl J Med. 2019;381(7):668–676. doi:10.1056/NEJMsr1809937

2. Bush K, Kivlahan DR, McDonell MB, Fihn SD, Bradley KA. The AUDIT alcohol consumption questions (AUDIT-C): an effective brief screening test for problem drinking. Arch Intern Med. 1998;158(16):1789–1795.

3. Bradley KA, Williams EC, Achtmeyer CE, Volpp B, Collins BJ, Kivlahan DR. Implementation of evidence-based alcohol screening in the Veterans Health Administration. Am J Manag Care. 2006;12(10):597–606.

4. Departments of Veterans Affairs and Defense (VA/DoD). VA/DoD Clinical Practice Guideline for the Management of Substance Use Disorders. Washington, DC: Department of Veterans Affairs, Department of Defense; 2015.

5. Office of the Inspector General, U.S. Department of Defense. Audit of Active Duty Service Member Alcohol Misuse Screening and Treatment (Report No. DODIG-2022-071). Alexandria, VA: Department of Defense; 2022.

6. Moore AA, Gould R, Reuben DB, et al. Longitudinal patterns and predictors of alcohol consumption in the United States. Am J Public Health. 2005;95(3):458–464.

7. Shao C, Wang X, Wang P, Tang H, He J, Wu N. Parkinson’s disease risk and alcohol intake: a systematic review and dose-response meta-analysis of prospective studies. Front Nutr. 2021;8:709846.

8. Letourneau B, Sobell LC, Sobell MB, Agrawal S, Gioia CJ. Two brief measures of alcohol use produce different results: AUDIT-C and Quick Drinking Screen. Alcohol Clin Exp Res. 2017;41(5):1035–1043. doi:10.1111/acer.13364

9. Rubinsky AD, Dawson DA, Williams EC, Kivlahan DR, Bradley KA. AUDIT-C scores as a scaled marker of mean daily drinking, alcohol use disorder severity, and probability of alcohol dependence in a U.S. general population sample of drinkers. Alcohol Clin Exp Res. 2013;37(8):1380–1390.

10. Cornell L, Shukla T, McMillan K, et al. Estimating the Causal Effect of Alcohol Consumption on Military-Relevant Mental Health Conditions: A Mendelian Randomization Study Using the All of Us Research Program. Manuscript submitted for publication. 2026.

11. Russell M, Welte JW, Barnes GM. Quantity-frequency measures of alcohol consumption: beverage-specific vs global questions. Br J Addict. 1991;86(4):409–417.

